# Impact of pre-existing T cell immunity to SARS-CoV-2 in uninfected individuals with COVID-19 mortality in different countries

**DOI:** 10.1101/2020.10.03.20206151

**Authors:** Gennadi V. Glinsky

**Affiliations:** Institute of Engineering in Medicine, University of California, San Diego, 9500 Gilman Dr. MC 0435, La Jolla, CA 92093-0435, USA

**Keywords:** COVID-19 pandemic, SARS-CoV-2, pre-existing reactive T cells, herd immunity, COVID-19 mortality

## Abstract

Several recent studies identified SARS-CoV-2 reactive T cells in people without exposure to the virus. However, pathophysiological implications of these findings remain unknown. Here, the potential impact of pre-existing T cell reactivity against SARS-CoV-2 in uninfected individuals on markedly different COVID-19 mortality levels in different countries has been investigated. The inverse correlation is documented between the prevalence of pre-existing SARS-CoV-2 reactive T cells in people without exposure to the virus and COVID-19 mortality rates in different countries. In countries with similar levels of pre-existing SARS-CoV-2 cross-reactive T cells in uninfected individuals, differences in COVID-19 mortality appear linked with the extend and consistency of implementations of social measures designed to limit the transmission of SARS-CoV-2 (lockdown; physical distancing; mask wearing). Collectively, these observations support the model that the level of pre-existing SARS-CoV-2 reactive T cells is one of the important determinants of the innate herd immunity against COVID-19. Together with the consistent social measures directed to limit the virus spread, high levels of pre-existing SARS-CoV-2 reactive T cells appear significant determinants diminishing the COVID-19 mortality. Observations reported in this contribution should have significant impact on definitions of the herd immunity threshold required to effectively stop the pandemic in different countries across the globe.

---

Several recent studies identified SARS-CoV-2 reactive T cells in uninfected individuals or people without exposure to the virus in various countries across the globe (1-5). It has been pointed out that the common major limitation of these studies was the relatively small numbers of donors contributing blood samples for the analyses (6). However, the most recent study (7) confirmed and extended these findings based on the analyses of 185 uninfected individuals, thus addressing to a significant degree this limitation. In unexposed to the SARS-CoV-2 individuals, the cross-reactive T cells were detected against 9 of 29 (31%) of the validated HLA class I and to 14 of 20 (70%) HLA-DR T cell peptide epitopes (7). Recognition frequencies against individual T cell epitopes in unexposed individuals were highly variable reaching up to 27% and 44% for the best-performing HLA class I and HLA-DR peptide epitopes, respectively (7). Therefore, studies utilizing validated individual peptide epitopes as well as predicted or random SARS-CoV-2-derived peptide pools, consistently demonstrated pre-existing SARS-CoV2-directed T cell responses in unexposed as well as seronegative for SARS-CoV-2 individuals (1-7). The likely source of this phenomena is the immune cross-reactivity between human common cold coronaviruses and SARS-CoV-2 (1-8).

Interestingly, the level of T cell reactivity against SARS-CoV-2 in unexposed individuals varied in different countries, ranging from 18% in Sweden (5) to 51% in Singapore (4). These findings prompted the analysis of the possible association between levels of T cell reactivity against SARS-CoV-2 in unexposed individuals and COVID-19 mortality rates in different countries. To this end, the currently available primary data on levels of T cell reactivity against SARS-CoV-2 in uninfected individuals and numbers of COVID-19 cases and death in five countries (United States; Netherlands; Germany; Singapore; Sweden) were obtained from original publications (1-5), analyzed (Methods), and plotted for visualization.

Notably, the significant inverse correlation (r = −0.991) has been observed between the levels of pre-existing T cell reactivity against SARS-CoV-2 and mortality rates in different countries (**Figure 1**). The most extreme cases were represented by Singapore, where 51% of population manifested pre-existing T cell reactivity associated with the COVID-19 mortality rate of 0.05%, compared with Sweden, where 18% of population manifested pre-existing T cell reactivity associated with the COVID-19 mortality rate of 6.35% (**Figure 1**).

**Figure 1.**
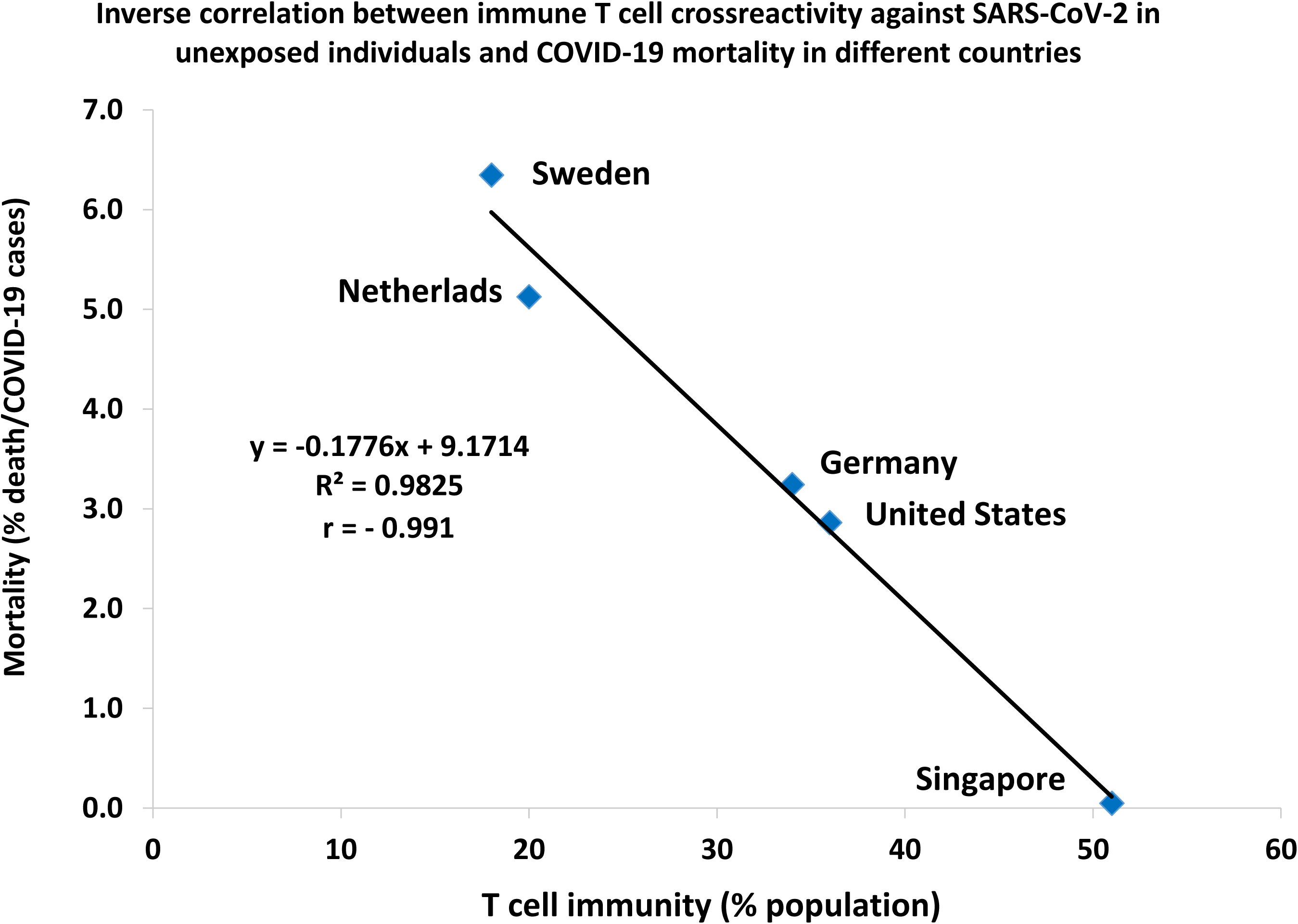
Inverse correlation between levels of pre-existing SARS-CoV-2 reactive T cells in uninfected individuals and COVID-19 mortality rates in different countries. COVID-19 mortality rates are reported as the case fatality rates (CFR) independently defined for each country (see Methods).

Interestingly, in two separate instances two countries manifesting similar levels of pre-existing T cell reactivity against SARS-CoV-2 in uninfected individuals reported markedly different COVID-19 mortality calculated as the numbers of death per million population (**Figure 2**). Germany and United States manifested similar levels of pre-existing T cell reactivity against SARS-CoV-2 in uninfected individuals: 34% and 36%, respectively. However, the reported COVID-19 mortality was 114 death per million in Germany and 627 death per million in United States. Similarly, Netherlands and Sweden manifested similar levels of pre-existing T cell reactivity against SARS-CoV-2 in uninfected individuals: 20% and 18%, respectively. However, the reported COVID-19 mortality was 374 death per million in Netherlands and 576 death per million in Sweden.

**Figure 2.**
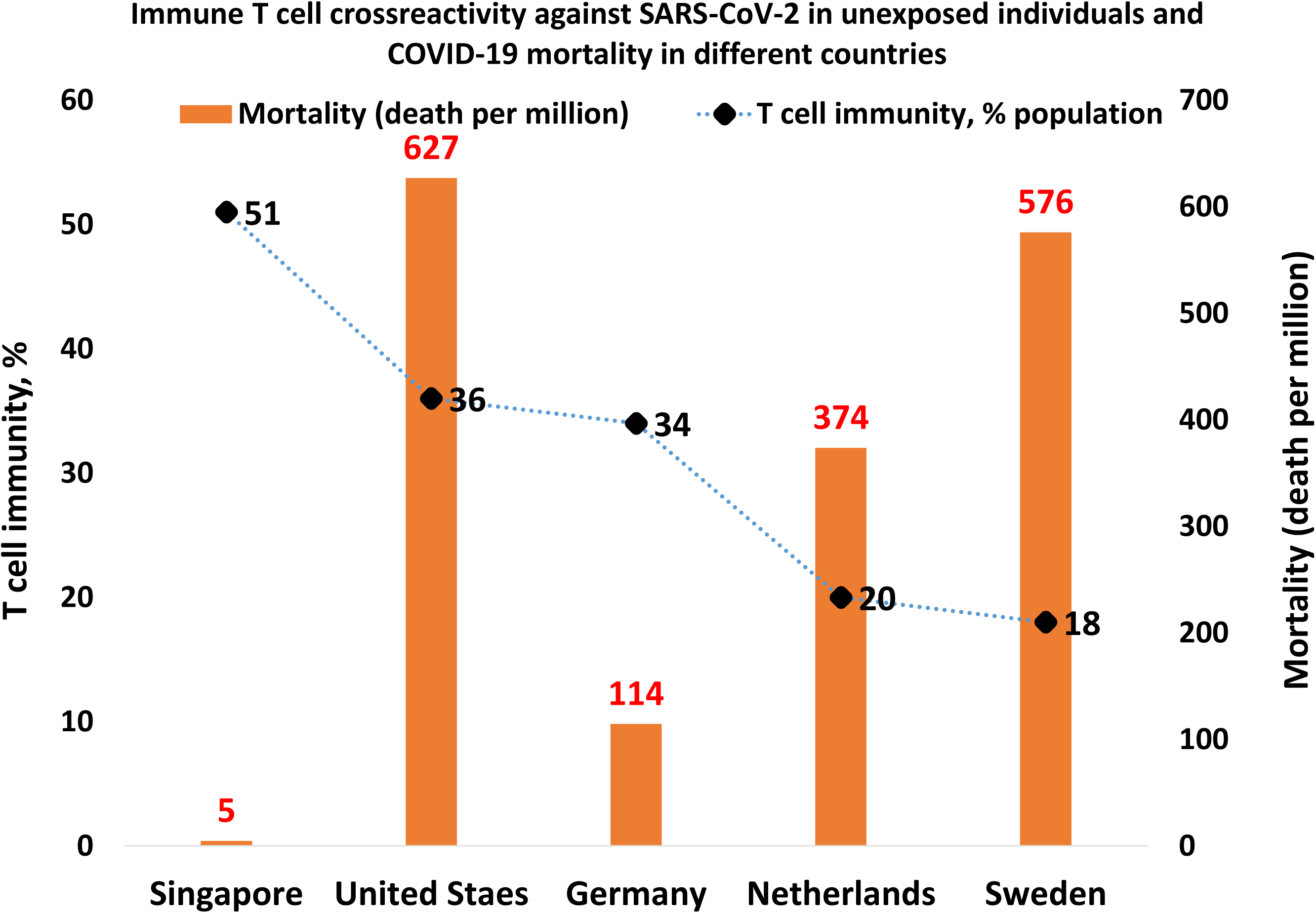
Countries with similar levels of pre-existing SARS-CoV-2 reactive T cells manifest different COVID-19 mortality normalized per million population.

Significantly, in both instances countries with lover COVID-19 mortality are known to adhere to more strict and consistent modes of implementations of social measures designed to limit the transmission of SARS-CoV-2 (lockdown; physical distancing; mask wearing) compared to their counterparts.

In contrast, no apparent association has been observed between the pre-existing T cell immunity against SARS-CoV-2 and numbers of COVID-19 cases in different countries (**Figure 3**). In both metrics reflecting either the mortality (**Figure 2**) or prevalence (**Figure 3**) of the COVID-19 pandemic, numbers reported for the United States appear notable outliers likely because systematic failures to articulate and consistently implement the national pandemic mitigation strategy designed to efficiently limit the spread of the virus.

**Figure 3.**
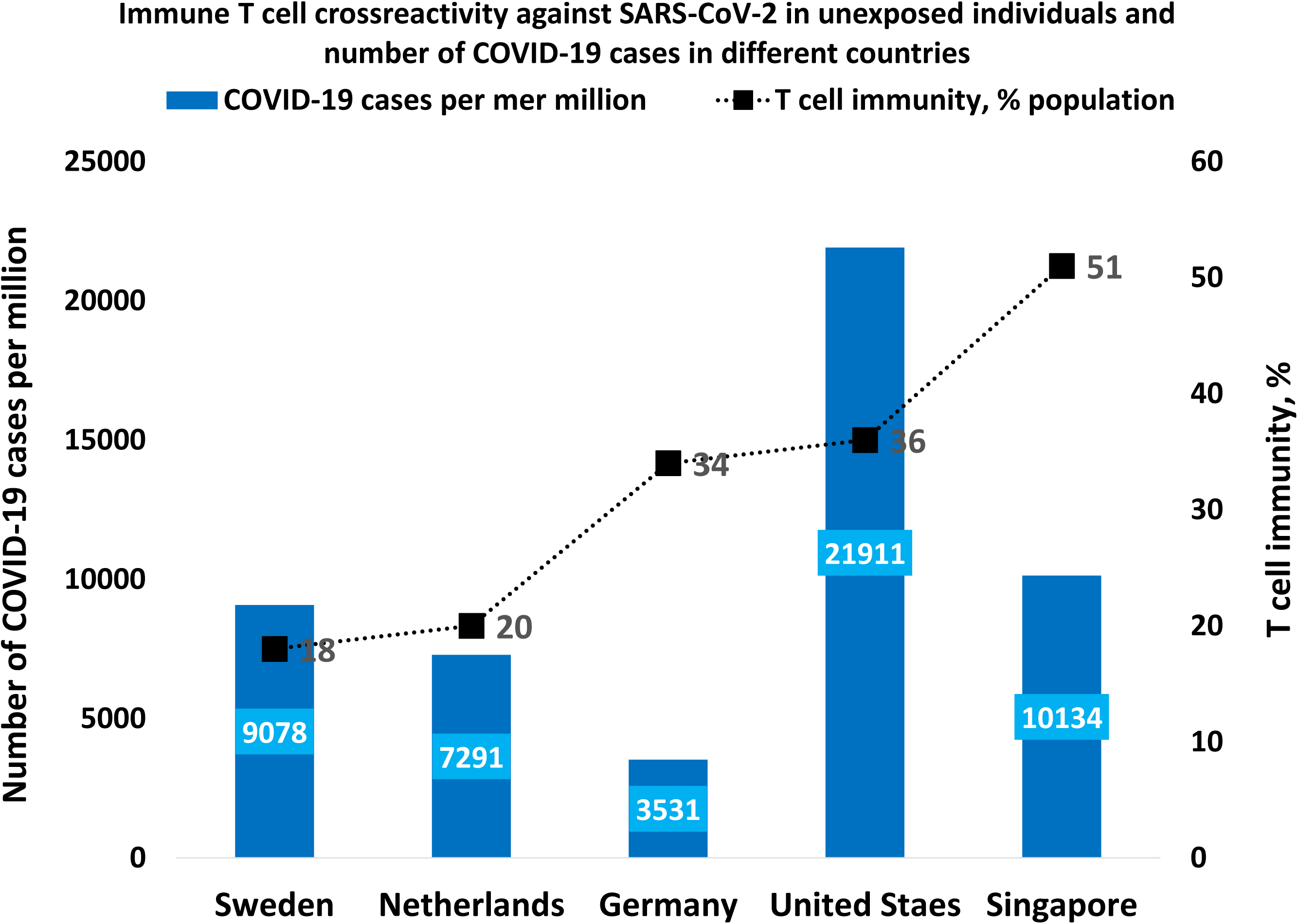
Lack of the apparent association between levels of pre-existing T cell reactivity against SARS-CoV-2 coronavirus in uninfected individuals and the prevalence of the COVID-19 pandemic in different countries. Prevalence of the pandemic was calculated as numbers of the reported COVID-19 cases normalized per million population.

Observations reported in this contribution indicate that pre-existing cross-reactive against SARS-CoV-2 T cells may contribute to the innate herd immunity against COVID-19 and affect populations’ susceptibility to the infection. Therefore, if pre-existing cross-reactive against SARS-CoV-2 T cells are significant contributors to populations’ herd immunity, the herd immunity threshold required to effectively stop the pandemic can be reduced from 60% of a population getting infected down to as low as 10% based on an R0 of 2.5 (the average number of secondary cases generated by an infectious individual among susceptible people) and achieved depending on the prevalence, quantity, and quality of pre-existing immunity among uninfected people (9-11). Based on these considerations, it seems likely that the upcoming implementation of even moderately efficient vaccination programs would facilitate the rapid attainment of the desirable herd immunity levels. If this conclusion is correct, we may be approaching the crossing of the herd immunity threshold required to stop the COVID-19 pandemic in not very distant future. Collectively, present findings indicate that the heterologous immunity facilitated by cross-reactive T cells pre-existing in uninfected individuals may be one of important contributing factors affecting the clinical course of the COVID-19 pandemic (7, 8) and strongly support the previously articulated rationale (3) for worldwide prospective studies precisely mapping the levels of pre-existing immune cross-reactivity against SARS-CoV-2 to the clinical course and outcomes of the pandemic.

## Limitations

One of the most significant limitations of this work is the small sample size of all reported to date studies on the levels of pre-existing T cell immunity against the SARS-CoV-2 in different countries, which is difficult to justify as representative samples of corresponding populations. Arguably, the COVID-19 mortality rate metric (also known as the case fatality rate, CFR) could be influenced by the lack of uniformity of diagnostic tests and the apparent inconsistency of the data collection and reporting in different countries, which could make uncertain the validity of observed differences between different countries. Future population-scale studies should help to address these uncertainties.

## Methods

The primary data on levels of T cell reactivity against SARS-CoV-2 in uninfected individuals are currently available for the following five countries: United States (1); Netherlands (2); Germany (3; 7); Singapore (4); Sweden (5). These data were obtained from original publications, supplemented with numbers of COVID-19 cases and death in corresponding countries (https://coronavirus.jhu.edu/map.html), analyzed, and plotted for visualization. Mortality rates were calculated for each country based on the statistics available on October 01, 2020 and reported as two metrics: i) percent of COVID-19 death normalized to number of COVID-19 cases (also known as case fatality rate, CFR); ii) number of COVID-19 death per one million population.

## Data Availability

All data referred to in the manuscript and note are available.

## Author Contributions

This is a single author contribution. All elements of this work, including the conception of ideas, formulation, and development of concepts, retrieval and analysis of data, and writing of the paper, were performed by the author.

## Acknowledgements

This work was made possible by the open public access policies of major grant funding agencies and international genomic databases and the willingness of many investigators worldwide to share their primary research data. This work was supported, in part, by OncoScar, Inc.

